# Association of Herpesvirus Response Burden with Long-Term Mortality Differs Between Older Males and Females

**DOI:** 10.1101/2020.11.12.20228916

**Authors:** Timo E Strandberg, Mikko Seppänen, Kaisu H Pitkälä, Mika Kivimäki, Pentti J Tienari

**Affiliations:** University of Helsinki and Helsinki University Hospital, Helsinki, Finland; University of Oulu, Center for Life Course Health Research, Oulu, Finland

**Keywords:** bacteria, herpes, immunoglobulin G, mortality, sex

## Abstract

**Background:** Sex-specific immune responses may contribute to variable vulnerability for Covid-19 between females and males. We tested whether there is a long-term mortality difference between sexes for other microbes (viral and bacterial) response burden among older people.

**Methods:** Seven-year follow-up study consisted of 382 home-dwelling people aged 75-90 years (65.2% females) with a history cardiovascular disease. At baseline, serum immunoglobulin G antibodies were assayed against herpesviruses (CMV, HSV-1 and HSV-2) and bacteria (*Chlamydophila pneumoniae, Mycoplasma pneumonia, and Helicobacter pylori*). Titers were summed up as herpes (HB) or bacterial response burden (BB) and divided into tertiles. Hazard ratios (HR) of total mortality with 95% CIs were calculated using Cox regression.

**Results:** The overall HB was lower and BB higher among males than females (P<0.001). There was a significant sex/HB (P=0.01) and sex/BB (P=0.03) interaction with mortality. Multivariable-adjusted (age, body mass index, C-reactive protein, and comorbidity index) mortality HRs for increasing HB sex-specific tertiles were 1.0 (reference), 1.34 (95% CI 0.62-2.88), and 2.66 (1.25-5.64) for males and 1.0, 1.30 (0.76-2.21), and 1.30 (0.77-2.22) for females. The significant age-adjusted association between BB and mortality in males attenuated after multivariable adjustments, HR (top-vs-bottom tertile) 1.74 (0.93-3.25). In females, no association with BB was observed. Using HB and BB as continuous variables supported the findings with tertiles.

**Conclusions:** Although being lower in older males than females, higher *Herpesviridae* response burden was associated with increased 7-year mortality risk among males, not among females. Immune responses to common microbes may contribute to sex differences in longevity and mortality.

**Key points:**

- Gender differences in vulnerability during Covid-19 has increased interested in sex-related responses to infections
- We used IgG titers of *Herpesviridae* and bacteria as surrogate markers for variably recurrent reactivation
- Although *Herpesviridae* response burden was generally lower among males than females, within sexes higher *Herpesviridae* burden strongly predicted 7-year mortality among males but not females
- Long-term virus burden, like *Herpesviridae*, may partly explain shorter longevity and higher mortality among males with weaker immune systems

## Introduction

There are evident sex differences in both innate and adaptive immune and inflammatory responses between sexes [1-3] and a greater proportion of males than females has been observed in the adverse clinical outcomes of SARS-CoV-2 disease (Covid-19) [4,5]. This male excess seems to be present in all age groups up to 80+ years of age in Europe [4]. Neutralizing autoantibodies against Type-1 interferons were recently discovered in patients with life-threatening Covid-19 pneumonia, 94% of the cases being males [6]. Type-1 interferon response is important in many viral infections, especially *Herpesviridae* [7]. Antibody titers against chronic pathogens are thought to reflect not only inter-individual variation in immunity but also reactivation frequency and thus could potentially act as surrogate markers for the frequency of reactivation and lack of infection control [8-10]. Typical common intracellular microbes with readily available serologic testing and tendency to prolonged or chronic infections include viruses in family *Herpesviridae* (herpes simplex virus types 1 [HSV-1] and [HSV-2] and cytomegalovirus [CMV]), and certain obligately (*Chlamydophila pneumoniae, Mycoplasma pneumoniae*) or facultatively (*Helicobacter pylori*) intracellular bacteria.

Since long-term mortality data of Covid-19 are not yet available, this prompted us to investigate whether sex differences in mortality would apply to other common infections. We tested associations between immunoglobulin G (IgG) antibody titers for HSV-1, HSV-2, CMV, *C. pneumoniae, M. pneumonia* and *H. pylori* and long-term mortality among persons aged ≥ 75 years, a group of people who are especially vulnerable to Covid-19 through numerous pathophysiological mechanisms related to ageing [11]. Besides being old and past reproductive age, our study subjects were community-dwelling and had a history of cardiovascular disease (CVD), which reduces the possibility that gender-patterned factors related to environment, lifestyle, current hormonal, and cardiovascular status would explain the findings.

## Methods

We performed secondary analyses of a prospective cohort study [12]. The cohort consists of individuals from the community, aged 75 to 90 years (mean 80 years) and with a history of stable CVD (n = 400, 65.2% females). They had participated in a multifactorial prevention trial between 2000 and 2003. Because the tested intervention did not affect mortality [12], all participants are handled as a single cohort. At baseline, they were investigated with questionnaires, clinical and laboratory examinations. As described earlier, information about lifestyle (body mass index [BMI], smoking), education (dichotomized as primary school only vs. higher), genetics (*APOE4* genotype), health-related quality of life (15D instrument [13]), morbidity, vitality and functional status (peak exploratory flow [PEF], renal function according to Chronic Kidney Disease Epidemiology Collaboration, CKD-EPI) [14], and Charlson comorbidity index [15]) was obtained. Serum C-reactive protein (CRP) and blood leukocytes were used to reflect inflammatory status.

At baseline in 2000, participants were also assayed for serum IgG antibodies against herpesviruses (CMV, HSV-1, HSV-2), *C. pneumoniae, M. pneumoniae* and *H. pylori* using enzyme immunoassay (EIA) methods, as described earlier [16,17]. Participants not tested for these biomarkers (N=18, 4.5% of those recruited) were excluded leading the analytical sample to consist of 382 participants. Clinically the excluded individuals did not differ from those included in the analysis. Absolute titers (U/mL) were used (except for *H pylori* for which titer was taken either 0 or 20 U/mL) to sum up *Herpesviridae* response burden (HB) or bacterial response burden (BB), which were used as surrogate markers for variably recurrent reactivation. In the analyses, HB and BB were used as continuous variables or divided in tertiles. Dichotomous seropositivity for each microbe was defined as described earlier [16,17].

Total mortality data and causes of death were retrieved from national register (Population Information System, Statistics Finland). Causes of death were divided in four main classes: coronary heart disease (CHD), other CVD, cancer, and other causes. Mortality during follow-up was investigated separately in males and females, and related to HB and BB at baseline. Because causes of death were available through 31 December, 2007, we restricted the follow-up time into seven years, during which 43.1% of the cohort (n=165) had died.

### Statistical analyses

The data were analyzed using NCSS statistical software (version 8, **www.ncss.com**) with descriptive statistics, analysis of covariance (ANCOVA), Kaplan–Meier plots, and Cox hazard models [hazard ratio (HR) with 95% confidence interval (CI)]. Two-sided *P* < 0.05 was considered statistically significant.

Cox regression was used to assess 7-year mortality risk and proportional hazard assumption was analyzed with Schoenfeld residuals. For the selection of adjusting variables, we investigated which clinically meaningful variables would predict total mortality in the whole cohort. The following variables were considered: age, PEF and BMI reflecting frailty, comorbidity index (dichotomous: >2 *vs* less) and CKD-EPI reflecting disease status, 15D sum score reflecting health related quality of life, *APOE4* genotype reflecting genetics, education reflecting social class, smoking history reflecting lifestyle, and log C-reactive protein reflecting inflammatory status. Of these variables, age, BMI, CRP, and comorbidity index independently predicted mortality in the whole cohort, and were used as covariates in the Cox analyses of HB, BB and mortality. Although MMSE reflecting cognition has been shown to predict mortality [18], it was not included here because it was also associated with microbial burden in this cohort [16].

### Ethics statement

All stages of the DEBATE study have been approved by the Ethics Committee of the Department of Medicine, Helsinki University Hospital, Helsinki, Finland. Informed consent according to the Declaration of Helsinki was obtained from all patients before any study procedures, which were performed according to good clinical trial practice (12).

## Results

Baseline characteristics of study participants are shown in Table 1. Females were on average slightly older, had higher BMI, and were more often never smokers than males. Females also had lower PEF, CKD-EPI, 15D sum score than males. There was no difference in C-reactive protein, leukocyte count and comorbidity index between sexes.

**Table 1.** Age-adjusted characteristics of females and males.

| Characteristic | Sex |  | P value between groups |
| --- | --- | --- | --- |
|  | Females, | Males, |  |
|  | n= 250 | n=132 |  |
| Age, yr, mean (SD) | 80.6 (0.3) | 79.1 (0.4) | 0.005 |
| - Median (interquartile range) | 80 (75-85) | 79 (75-80) |  |
| Primary school only, n (%), | 98 (39.5) | 43 (32.8) | 0.20 |
| Never smoker, n (%) | 173 (69.2) | 41 (31.1) | <0.001 |
| APOE4, n (%) | 75 (30.9) | 37 (28.0) | 0.57 |
| Body-mass index, kg/m<sup>2</sup> | 26.9 (0.3) | 25.9 (0.4) | 0.019 |
| MMSE, mean (SD) | 26.2 (0.2) | 26.5 (0.2) | 0.40 |
| C-reactive protein, mg/L | 3.1 (0.3) | 2.9 (0.5) | 0.45 (log) |
| Glucose, mmol/L | 5.4 (0.08) | 5.3 (0.1) | 0.59 |
| Leucocyte count, | 6.4 (0.1) | 6.5 (0.1) | 0.09 (log) |
| CKD-EPI, mL/min/1.73 m<sup>2</sup> | 54.1 (0.7) | 58.6 (1.0) | <0.001 |
| Peak expiratory flow, mL/min | 304.1 (5.0) | 407.3 (6.8) | <0.001 |
| 15D score (0=worst, 1.0=best) | 0.78 (0.01) | 0.81 (0.01) | 0.006 |
| Comorbidity index <sup>a</sup> | 2.5 (0.1) | 2.3 (0.1) | 0.26 |

**Table 1 continued**
|  |  |  |  |
| --- | --- | --- | --- |
| HS V-1 IgG, U/mL | 105.8 (3.2) | 115.1 (4.4) | 0.09 |
| -seropositive, % | 88.4 | 88.6 | 0.95 |
| HS V- 2 IgG, U/mL | 53.9 (3.8) | 26.2 (5.2) | <0.001 |
| -seropositive, % | 42.8 | 18.2 | <0.001 |
| CMV IgG, U/mL | 128.0 (6.8) | 88.2 (9.3) | <0.001 |
| -seropositive, % | 92.4 | 87.9 | 0.14 |
| ***Herpesviridae* response burden <sup>b</sup>,** | 287.8 (8.2) | 229.6 (11.3) | <0.001 |
| IgG, U/mL |  |  |  |
| *C pneumonia* IgG, U/mL | 84.1 (6.1) | 132.1 (8.4) | <0.001 |
| -seropositive, % | 56.3 | 77.4 | <0.001 |
| *M pneumoniae* IgG, U/mL | 65.8 (6.0) | 83.0 (8.3) | 0.09 |
| *H pylori* IgG, <sup>c</sup> U/mL | .. | .. |  |
| -seropositive, % | 55.2 | 58.3 | 0.55 |
| **Bacterial response burden <sup>d</sup>** | 160.8 (9.1) | 226.9 (12.5) | <0.001 |
| U/mL |  |  |  |

Continuous variables are age-adjusted mean (SE) unless otherwise indicated. Analysis of covariance for continuous variables. CKD-EPI = Chronic Kidney Disease Epidemiology Collaboration; CMV= cytomegalovirus; HSV = herpesvirus. <sup>a</sup> Calculated according to Charlson et al [15]. <sup>b</sup>Sum of HSV-1, HSV-2, and CMV IgG levels. <sup>c</sup> Taken as seronegative = 0, seropositive = 20 U/mL. <sup>d</sup> Sum of *C pneumonia*, *M pneumoniae*, and *H pylori* IgG levels

Of different IgG antibody levels, females had higher CMV and especially HSV-2 titers whereas HSV-1 titers were similar leading to significantly higher overall HB among females. In contrast, males had constantly higher anti-bacteria titers and consequently higher BB.

Cumulative mortality during 7 years was similar between sexes (44.6%, n=62 in males and 39.5%, n=103, among females, P=0.32). However, after adjusting for age males had 42% higher mortality than females (HR 1.42, 95% CI 1.03-1.96). Overall, 34.0%, 21.6%, 17.0%, and 25.5% of deaths were due to CHD, other CVD, cancer and other causes, respectively (cause of death was unknown in 2.0%). No significant differences in these causes of death were observed between sexes, nor between HB and BB tertiles.

As shown in Figure, unadjusted, sex-specific Kaplan-Meier curves revealed a significant mortality difference between HB tertiles in males (P=0.048), but not among females. For BB tertiles of males, mortality difference tended to be significant (P=0.061). Further analyses showed that there was a significant sex x HB (P=0.01) and sex x BB (P=0.03) interaction with mortality. We also analyzed the association of individual microbes to mortality. For HB, the association was mainly driven by CMV, for BB no differences between bacteria were found (data not shown).

In age-adjusted Cox analyses among females, mortality HRs for increasing HB sex-specific tertiles were 1.0 (reference), 1.17 (95% CI 0.70, 1.97), and 1.30 (0.78, 2.18) (Table 2). The corresponding HRs among males were 1.0 (reference), 1.21 (0.62, 2.60), and 2.14 (1.10, 4.18), respectively. The results remained after adjustment for BMI, C-reactive protein, and comorbidity index (among males HR in the top vs bottom tertile 2.66, 95% CI 1.25-5.64).

**Table 2.** Serially adjusted Cox regression for 7-year mortality according to baseline microbial response burden in males and females.

| **Hazard ratio (95% CI) for 7-year total mortality** |  |  |  |  |  |  |
| --- | --- | --- | --- | --- | --- | --- |
| *Herpesviridae* response burden tertiles<sup>a,c</sup> |  |  |  |  |  |  |
| Model | Women |  |  | Men |  |  |
|  | 1 | 2 | 3 | 1 | 2 | 3 |
| Unadjusted | 1.0 | 1.24 | 1.60 | 1.0 | 1.36 | 2.19 |
|  | (reference) | (0.74-2.08) | (0.96-2.65) | (reference) | (0.67-2.79) | (1.12-4.27) |
| Adjusted for age | 1.0 | 1.17 | 1.30 | 1.0 | 1.21 | 2.14 |
|  | (reference) | (0.70-1.97) | (0.77-2.18) | (reference) | (0.59-2.47) | (1.10-4.18) |
| Adjusted for age, log CRP, BMI, and comorbidity index<sup>b</sup> | 1.0 | 1.30 | 1.30 | 1.0 | 1.34 | 2.66 |
|  | (reference) | (0.76-2.21) | (0.77-2.22) | (reference) | (0.62-2.88) | (1.25-5.64) |
| **Bacterial response burden tertiles<sup>a,d</sup>** |  |  |  |  |  |  |
| Model | Women |  |  | Men |  |  |
|  | 1 | 2 | 3 | 1 | 2 | 3 |
| Unadjusted | 1.0 | 0.92 | 0.72 | 1.0 | 0.96 | 1.83 |
|  | (reference) | (0.58-1.46) | (0.43-1.21) | (reference) | (0.48-1.96) | (0.99-3.38) |
| Adjusted for age | 1.0 | 0.84 | 0.70 | 1.0 | 1.10 | 1.94 |
|  | (reference) | (0.53-1.33) | (0.42-1.18) | (reference) | (0.54-2.23) | (1.05-3.58) |
| Adjusted for age, log CRP, BMI, and comorbidity index<sup>b</sup> | 1.0 | 0.84 | 0.63 | 1.0 | 1.42 | 1.74 |
|  | (reference) | (0.52-1.37) | (0.38-1.05) | (reference) | (0.67-3.02) | (0.93-3.25) |

BMI= body mass index, CRP= C-reactive protein. <sup>a</sup>Sex-specific tertiles. <sup>b</sup>Selection of adjusting variables, please see Methods. <sup>c</sup> *Herpesviridae* response burden consisted of the sum of serum immunoglobulin G antibodies against herpes simplex virus type 1 (HSV-1), herpes simplex virus type 2 (HSV-2), and cytomegalovirus (CMV), divided in tertiles. <sup>d</sup> Bacterial response burden consisted of the sum of serum immunoglobulin G antibodies against *Chlamydia pneumoniae*, *Mycoplasma pneumonia*, and *Helicobacter pylori*.

With BB, the significant age-adjusted association with mortality in males (HR 1.94, 95% CI 1.05-3.58) attenuated after multivariable adjustment (1.74, 95% CI 0.93-3.25) (Table 2). Among females, the association was inverted, but not statistically significant.

Associations of HB and BB with total mortality were also examined as continuous variables and adjusted for age, comorbidity index, BMI, and log C-reactive protein. Again, HB predicted mortality among males (HR per SD increase 2.02, 95% CI 1.46-2.80), but not among females (HR per SD increase 1.16 (95% CI 0.96-1.41). For BB, HR per SD was 1.23 (95% CI 0.96-1.59) for males and 0.86 (95% CI 0.68-1.09) for females.

## Discussion

While *Herpesviridae* response burden, defined as summed antibody titers of HSV-1, HSV-2 and CMV was higher among 75+ females than males, increasing HB was significantly associated with long-term mortality only among males. Although tentative among males, similar associations were nonsignificant for BB defined as summed titers for *C pneumoniae, M pneumoniae* and *H pylori*. All study participants were home dwelling but with a history of CVD, that is patient group shown to be vulnerable for coronavirus infection.

Males and females are known to have different risks and responses to various autoimmune and infectious conditions as well as differences in the regulation of systemic inflammation [1-3,19,20], but the sex difference has been rarely taken into account in analyses [1]. Recently, the Covid-19 disease has attracted research in the sex and gender biases, because male patients are more vulnerable to Covid-19 [4,5]. One factor underlying male vulnerability was recently identified, the neutralizing antibodies against type 1 interferons (6). This finding is highly relevant in light of the present study since type 1 interferons have been identified as major factors in host defense against *Herpesviridiae* [7]. Differences in immune responses due to genes (XY vs XX karyotypes), epigenetics, hormones and gender-specific behavioral factors may explain sex differences in longevity and mortality.

The relationship between immune response and outcome has been described as a U-curve [21]. Adaptive immunity is important for the control of chronic intracellular infections, CD8+ T cells being the major responsible cell type. Immunosenescence, *i*.*e*. the effect of ageing on the immune system, is more prominent in males as indicated by the number of senescent CD8+ T cells and their ratio to total CD8+ T cell counts [22]. Excess damage may occur both due to too weak and too strong immune responses. Lower overall antibody titer among men and higher among women might then partly explain the observed mortality difference. Male bias may be seen when weaker male immune responses – insufficient to resist microbes – are associated with higher levels of host damage [1]. Female bias, in turn, is observed when strong immune response contributes to host damage and autoimmunity [1]. Efficiency of T cell responses is inversely associated with age and a poor T cell response is related to worse disease outcome in males but not in females [23]. Plausible explanations could include the effect of other gender differences: males have more risk factors like smoking, poor oral health, and unhealthy lifestyles with consequent mild chronic inflammation contributing to cardiovascular diseases [24]. Therefore, males may also be more prone than females to smoldering endotheliitis and endothelial dysfunction, a suggested key mechanism in the pathogenesis of Covid-19 [25].

However, our results in an older cohort with similar morbidity status and history of CVD suggest that the sex difference may apply to viral infections in general, and be independent of common inflammatory markers and of gender differences due to comorbidity or vascular function. Because all our participants were well past reproductive age, it is unlikely that female or male hormonal factors known to affect immunity (1) would offer a plausible explanation. Genetic factors were not extensively studied, but *APOE4* status, one of the few genetic markers known to affect mortality, longevity and herpesvirus susceptibility in humans [26,27], did not explain sex differences for HB.

Why was HB more strongly than BB related to mortality? Pathogens causing BB are treatable, less prolonged or chronic. One of the answers may relate to the age distribution of our older participants. When the study subjects were young, *Herpesviridae* like CMV and HSV-1 were frequently acquired early in life. Antibody levels probably reflect life-long, persistent immunologic burden and the response to it. Higher CMV titers are related to the above-mentioned CD8+ T cell immunosenescence [28]. HB may thus stress the weaker, immunologically frail male system more profoundly than a potentially shorter-term and transient BB. Considering Covid-19, it is of interest whether worse prognosis in males is associated, in addition to Type-1 interferon autoantibodies [6], with greater chronic HB or otherwise more advanced CD8+ T-cell immunosenescence. In recent analyses, antibody responses to viruses, including HSV-1 and CMV, have been reported to be associated with Covid-19 severity [29].

## Strengths and limitations

Limitations of our study include that participants are selected, relatively well-performing home-dwelling individuals aged 75 years and older and with stable CVD. On the other hand, the participants were drawn from a population-based sample and similar CVD background may be a strength to control for characteristics otherwise different among men and women and contributing to mortality. Immune status was assessed with antibody titers at baseline and more sophisticated methods of immune cells or direct measurements of microbial load were not available. Therefore, we can only speculate if for example higher titers among women *vs* lower titers among men were due to activated or suppressed response, respectively, or due to stable differences between sexes. Crude indicators of innate immunity, C-reactive protein level and blood leukocytes, were, nevertheless, similar between males and females. Neither were there signs that males would have been frailer than females, rather the opposite: although comorbidity indices were similar, PEF and health-related quality of life were significantly better among men who were also slightly younger than the women. Finally, because of their age (75 years and older), observed sex differences can be less attributable to the effects of sex hormones.

## Conclusion

Long-term mortality was strongly associated with *Herpesviridae* antibody response burden among older males, but not among females. Male bias during the ongoing Covid-19 pandemia may thus reflect a specific difference in susceptibility to the virus or deleterious effects due to certain other viral infections such as *Herpesviridiae*. This male bias may further contribute to longevity differences between sexes and should be duly taken into account in prevention and treatment of viral diseases and men’s health [30].

## Data Availability

Available on request

## Legend to the Figure

### Crude cumulative mortality in males and females according to *Herpesviridae* and bacterial response burden

*Herpesviridae* response burden (HB) consisted of the sum of serum immunoglobulin G antibodies against herpes simplex virus type 1 (HSV-1), herpes simplex virus type 2 (HSV-2), and cytomegalovirus (CMV), divided in tertiles. Bacterial response burden (BB) consisted of the sum of serum immunoglobulin G antibodies against *Chlamydia pneumoniae, Mycoplasma pneumoniae* and *Helicobacter pylori* divided in tertiles.

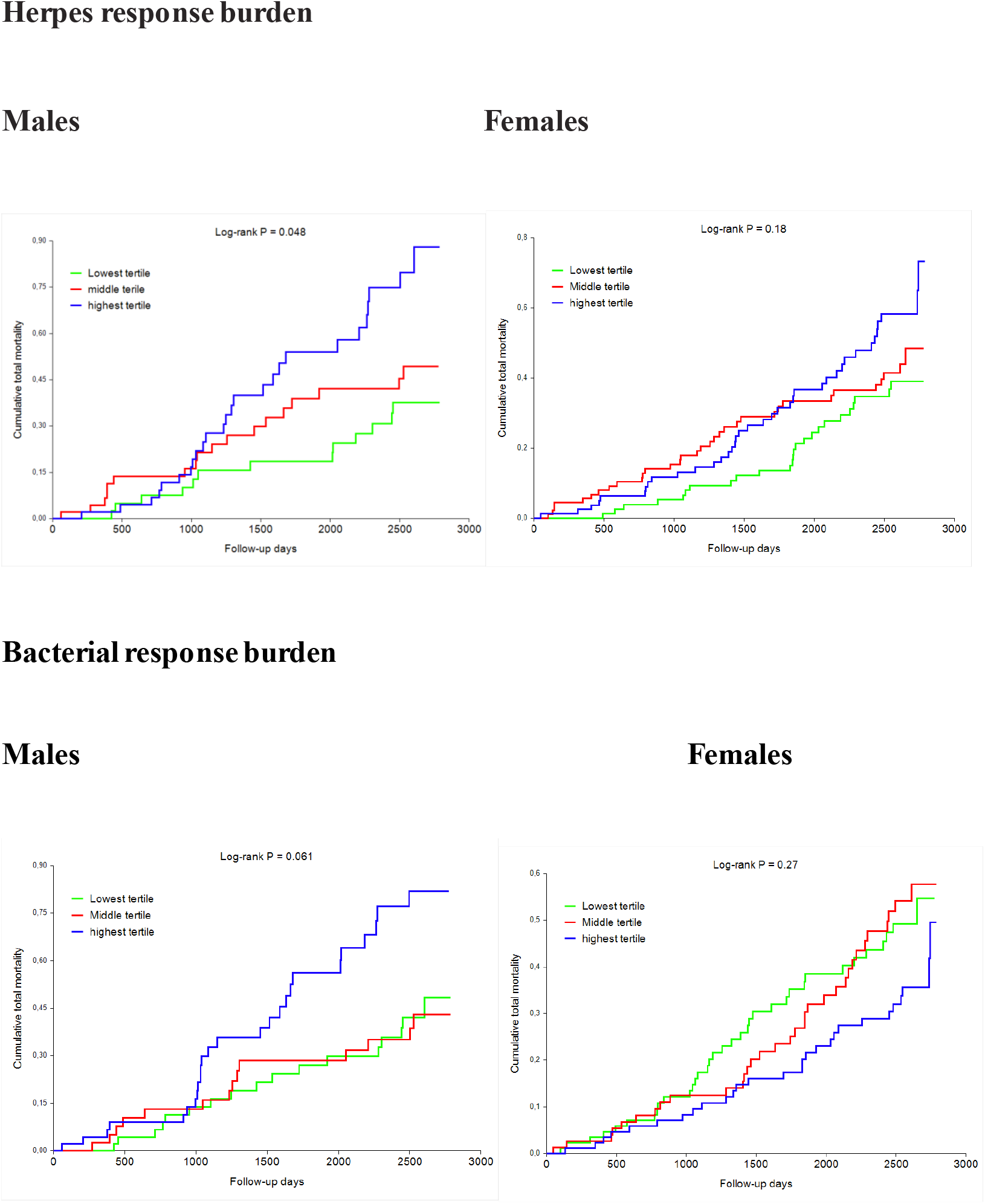

